# Age-stratified Prevalence and Relative Prognostic Significance of Traditional Atherosclerotic Risk Factors: A Report from the Nationwide Registry of Percutaneous Coronary Interventions in Japan

**DOI:** 10.1101/2023.05.02.23289427

**Authors:** Kenji Kanenawa, Kyohei Yamaji, Shun Kohsaka, Hideki Ishii, Tetsuya Amano, Kenji Andò, Ken Kozuma

## Abstract

**Background:** The prevalence of traditional atherosclerotic risk factors (TARFs) and their association with clinical profiles or mortality in percutaneous coronary intervention (PCI) remain unclear.

**Methods:** The study analyzed 559,452 patients who underwent initial PCI between 2012 and 2019 in Japan. TARFs were defined male, hypertension, dyslipidemia, diabetes, smoking, and chronic kidney disease (CKD). We evaluated the age-specific prevalence of TARFs, and calculated the relative importance (RI) according to R^2^, machine learning to assess the impact of TARFs on clinical profile (age, the presentation of acute myocardial infarction, cardiogenic shock, and multivessel disease) and in-hospital mortality. The average percentage of the RI calculated from these models was defined as the relative contribution (RC) of each TARF.

**Results:** The age-specific prevalence of TARFs, except for CKD, formed an inverted U-shape with significantly different peaks and percentages. Smoking was more prevalent in patients with ST-elevation myocardial infarction than in patients with stable angina (odds ratio [OR], 1.75, 95% confidence interval [CI], 1.73-1.77). In logistic regression model and relative risk model, smoking was most strongly associated with acute myocardial infarction (AMI) (adjusted OR, 1.62, 95% CI, 1.60-1.64; RC, 47.1%) and premature coronary artery disease (adjusted unstandardized beta coefficient [UC], 2.68, 95% CI, 2.65-2.71, RC, 42.2%). Diabetes was most strongly associated with multivessel disease (adjusted UC, 0.068; 95% CI, 0.066-0.070, RC, 59.4%) and the presentation of non-AMI (adjusted OR, 0.77, 95% CI, 0.76-0.78, RI, 21.9%). The absence of dyslipidemia was most strongly associated with presentation of cardiogenic shock (adjusted OR, 0.62; 95% CI, 0.61-0.64, RC, 34.2%) and in-hospital mortality (adjusted OR, 0.44, 95% CI, 0.41-0.46, RC, 39.8%). These specific associations were consistently observed regardless of adjustment or stratification by age.

**Conclusions:** Our analysis showed a significant variation in the age-specific prevalence of TARFs. Further, their contribution to clinical profiles and mortality also varied widely.

**What is known:**

- Traditional atherosclerotic risk factors (TARFs) are well-established targets for the prevention of adverse cardiovascular events.
- The impact of TARFs on clinical profiles in the patients with percutaneous coronary intervention remain unclear.

**What the Study Adds:**

- Due to the prevalence of TARF, which varies by age group, the TARFs that should be actively intervened and educated on will differ for each age group
- The relative importance of TARF differed significantly by clinical profile (age, the presentation of acute myocardial infarction, cardiogenic shock, and multivessel disease) and in-hospital mortality.
- Understanding the differences in the impact of TARFs on CAD is an important step in improving population-based strategies for CAD prevention.

## Introduction

Coronary artery disease (CAD), remains a leading cause of death in developed countries, with various genetic and environmental risk factors contributing to its development. Among these, traditional atherosclerotic risk factors (TARFs), established by epidemiological cohort studies decades ago, continue to play a crucial role in CAD prevention and management ^1 2^. Recent studies have demonstrated that even in the contemporary era, 80-85% of CAD patients have at least one TARF ^3–6^. The management of these risk factors has led to a continuous reduction in the risk of future cardiovascular events^7^.

In addition to their direct effects on atherosclerosis development, TARFs have been shown to contribute to the development of comorbidities such as heart failure^8^, which in turn may influence intermediate pathways related to mortality. However, the impact of TARFs on the clinical profile or mortality of patients who undergo revascularization procedures, including percutaneous coronary intervention (PCI) remain uncertain. Understanding the differences in its age-specific prevalence and the impact of TARFs on CAD is an important step in improving population-based strategies for CAD prevention, as well as promoting equitable evidence-based treatment during revascularization.

To this end, we evaluated the strength of the association of TARF with clinical profiles and mortality rates using data from a prospective multicenter nationwide PCI registry. Our findings may have important implications for improving risk stratification and management of patients with CAD, as well as for designing effective population-based prevention strategies.

## Methods

### Study design and ethical approval

The Japanese Percutaneous Coronary Intervention (J-PCI) registry is an ongoing prospective multicenter nationwide PCI registry maintained by the Japanese Association of Cardiovascular Intervention and Therapeutics^9, 10^. In addition, the J-PCI registry has been incorporated into the National Clinical Data System, which is a nationwide prospective web-based registry linked to medical and surgical board certification since 2013 ^11^.Cardiac catheterization procedures are performed in both public and private hospitals in Japan, and because registration in the J-PCI database is mandatory for both systems, data completeness is high. Compared with the annual reports from the insurance claims data, approximately 90% of all PCI procedures in Japan were registered in the J-PCI registry^7^. The J-PCI study protocol conformed to the principles outlined in the Declaration of Helsinki and was approved by the Institutional Review Board of the Network for Promotion of Clinical Studies (a specialized nonprofit organization affiliated with the Osaka University Graduate School of Medicine in Osaka, Japan). As all data were anonymous and aggregated without any personal information, ethical approval was waived.

The present study included patients who underwent initial PCI from January 2012 to December 2019 in the J-PCI Registry. Patients with missing data regarding baseline characteristics and/or those aged < 18 or > 100 were excluded. Additionally, we excluded patients with a history of CAD and/or any coronary revascularization to evaluate the onset age of CAD in patients with or without each TARF.

### Variable definitions

In this study, we defined seven factors, including male sex, smoking, hypertension, dyslipidemia, diabetes, and chronic kidney disease (CKD), as TARFs^12–14^. The exact definitions of risk factors in the J-PCI registry are described elsewhere^15^. Briefly, smoking was defined as any history of smoking within the past year. Hypertension was defined as systolic blood pressure ≥ 140 mmHg, diastolic blood pressure ≥ 90 mmHg, or the use of antihypertensive medications. Dyslipidemia was defined as fasting triglyceride levels ≥ 150 mg/dL, fasting high-density lipoprotein cholesterol levels < 40 mg/dL, fasting low-density lipoprotein cholesterol levels calculated using the Friedewald equation ≥ 140 mg/dL, non-high-density lipoprotein cholesterol levels ≥ 170 mg/dL, or the use of anti-hyperlipidemic medications. Diabetes mellitus was defined as fasting plasma sugar levels ≥ 126 mg/dL, random plasma sugar levels ≥ 200 mg/dL, hemoglobin A1c levels ≥ 6.5%, plasma glucose levels 2 h after a 75-g oral glucose tolerance test ≥ 200 mg/dL, or the use of anti-diabetic medications. CKD was defined as the presence of proteinuria, a serum creatinine level ≥ 1.3 mg/dL, or an estimated glomerular filtration rate ≤ 60 mL/min per 1.73 m^2^. In this study, end-stage renal failure with dialysis was included in the definition of CKD.

### Statistical analysis

Continuous variables were expressed as mean ± standard deviation or as median (interquartile ranges) and were compared using Student’s *t*-test or Wilcoxon rank-sum test. Categorical variables were presented as values and percentages and were compared using the χ^2^ test. Differences in baseline characteristics were evaluated based on the presence or absence of each TARF. We calculated odds ratios (OR) for the prevalence of TARFs by four CAD types: ST-elevated myocardial infarction (STEMI), non-STEMI (NSTEMI), unstable angina (UA), and stable angina (SA). The prevalence of TARF by age and CAD types were estimated using a logistic regression model with a cubic spline function. Patients aged 20-29 years were excluded from cubic spline function because of their small number and the wide variation in the prevalence of TARF.

To evaluate the strength of the association of TARFs on clinical profiles (age at CAD onset requiring PCI, multivessel disease, the presentation of STEMI, and cardiogenic shock) in a cross-sectional study approach, we used the unstandardized beta coefficient in multiple regression analysis, or the OR derived from multivariate logistic analysis. Similarly, the association between in-hospital mortality and TARFs was evaluated. Then, we estimated the relative importance (RI) among TARFs by R^2^ in multiple regression analysis or multivariate logistic regression analysis. For the R^2^ value in the multivariate logistic regression analysis, we used McFadden’s pseudo-R^2^ value. In addition, the RI was also calculated by Gain of gradient boosting and Mean Decrease Accuracy (MDA) of random forests to corroborate these findings. The unit for RI according to gradient boosting and random forests was arbitrary and was normalized to 100 to facilitate interpretation. The RI of each TARF from the three models was converted into percentage units as relative contribution (RC). For example, when the R^2^ value for smoking was 0.01 and the sum of R^2^ for all variables was 0.1 in logistic regression model, the RC of smoking was interpreted as 10.0% (0.01/0.1). As noted above, the RC obtained from the machine learning models can also be treated as a percentage of 100, so the average RC of each TARF from the three models was used to evaluate the strength of their associations with clinical profile and outcome.

A two-sided P value < 0.05 was considered statistically significant for all tests. All analyses were performed using the R software (version 4.1.1; R Foundation for Statistical Computing, Vienna, Austria).

## Results

### Baseline characteristics

This study included a total of 559,452 patients who underwent their first PCI at 1,123 centers covered by the registry. The mean age of patients was 70.0 ± 11.6 years, and the male sex accounted for 73.3%. The age-specific prevalence of the TARFs is shown in **Figure 1**. The prevalence of male sex, hypertension, dyslipidemia, smoking, and diabetes exhibited an inverted U-shaped curve, while their peaks differed significantly. The peak prevalence of male sex, smoking, and dyslipidemia was in younger patients, that of diabetes was in middle-aged patients, and that of hypertension and CKD was in older patients. The prevalence of CKD increased steadily and sharply with age.

**Figure 1.**
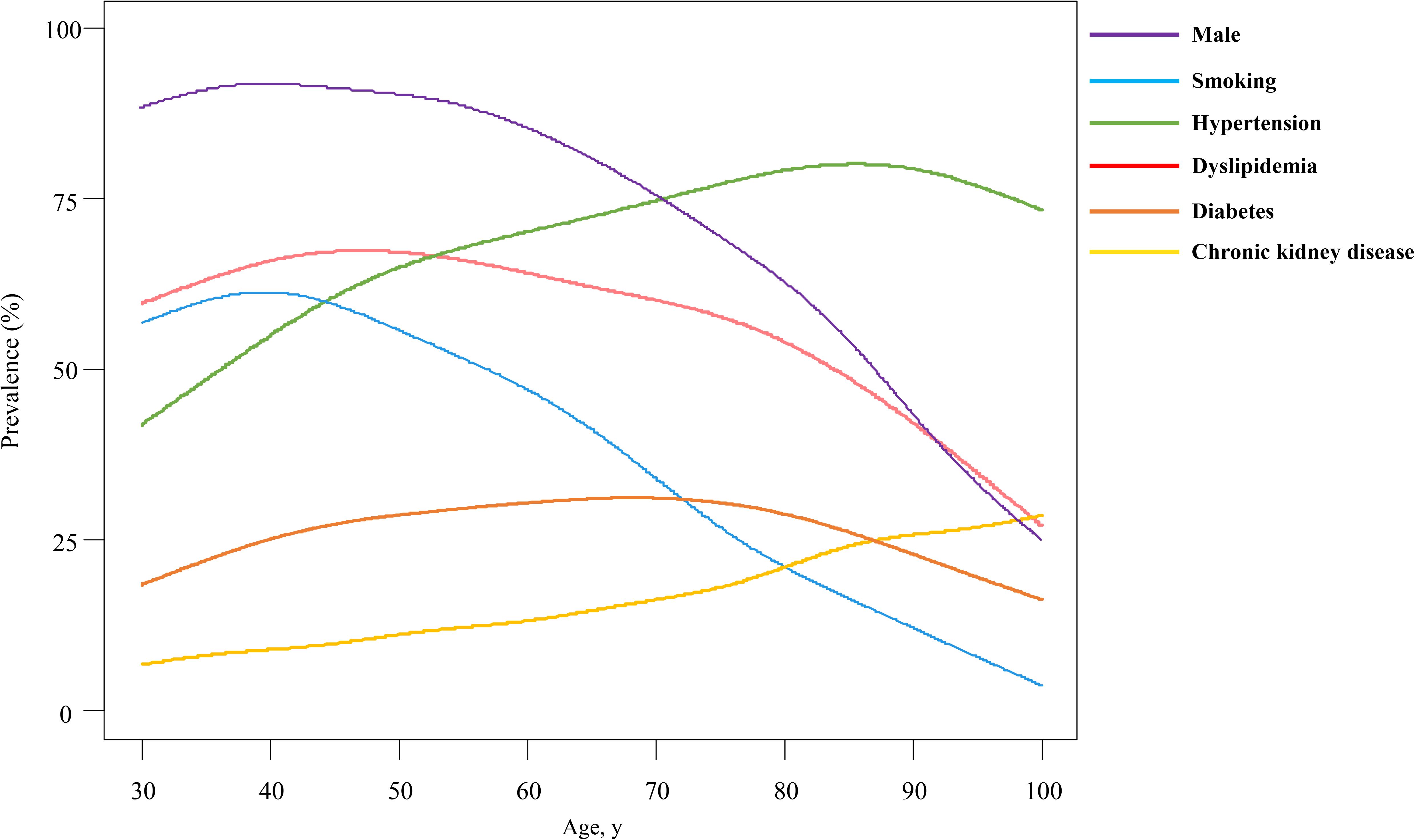
Age-specific prevalence of traditional atherosclerotic risk factors with a cubic spline.

The prevalence of TARFs by CAD type were evaluated with reference to SA (**Table 1**). The prevalence of smoking in patients with STEMI relative to those with SA was significantly higher (OR, 1.75, 95% Confidence interval [CI], 1.73-1.77). On the other hand, the prevalence of hypertension (OR, 0.62, 95%CI, 0.61-0.62), dyslipidemia (OR, 0.77, 95%CI, 0.76-0.78), diabetes (OR, 0.66, 95%CI, 0.65-0.67), and CKD (OR, 0.69, 95%CI, 0.68-0.70) in patients with STEMI were lower than those with SA. The prevalence of STEMI was U-shaped with a peak in the 70s and inversely correlated with the overall prevalence of AP (**Figure 2**). It was noteworthy that the prevalence of STEMI was higher among young smokers and that the prevalence of AP was higher among young diabetic patients.

**Table 1.** Prevalence of traditional risk factors by clinical presentations.

|  | Male | Smoking | Hypertension | Dyslipidemia | Diabetes | Chronic kidney disease |
| --- | --- | --- | --- | --- | --- | --- |
| Stable angina | 72.2% (Reference) | 27.4% (Reference) | 74.8% (Reference) | 60.4% (Reference) | 42.4% (Reference) | 16.5% (Reference) |
| ST-elevation myocardial infarction | 74.9% (1.17 [1.15-1.18]) | 40.0% (1.75 [1.73-1.77]) | 64.7% (0.62[0.61-0.62]) | 54.1% (0.77 [0.76-0.78]) | 32.8% (0.66 [0.65-0.67]) | 12.0% (0.69 [0.68-0.70]) |
| Non-ST-elevation myocardial infarction | 75.2% (1.15 [1.12-1.18]) | 37.2% (1.57 [1.53-1.60]) | 69.8% (0.78[0.76-0.80]) | 57.4% (0.88 [0.86-0.90]) | 35.4% (0.75 [0.73-0.76]) | 14.9% (0.88 [0.86-0.91]) |
| Unstable angina | 72.3% (1.01 [0.99-1.02]) * | 31.8% (1.23 [1.21-1.25]) | 71.0% (0.82[0.81-0.84]) | 60.1% (0.99 [0.97-1.00]) * | 35.6% (0.75 [0.74-0.76]) | 12.9% (0.75 [0.73-0.76]) |

Categorical variables are given as percentage (odds ratio [95% CI]). Odds ratios were calculated with reference to stable angina. The comparison between presence and absent of each atherosclerotic risk factor are significant at P
<0.001 unless otherwise noted.
\*The comparison between presence and absence of each atherosclerotic risk factor are nonsignificant.

**Figure 2.**
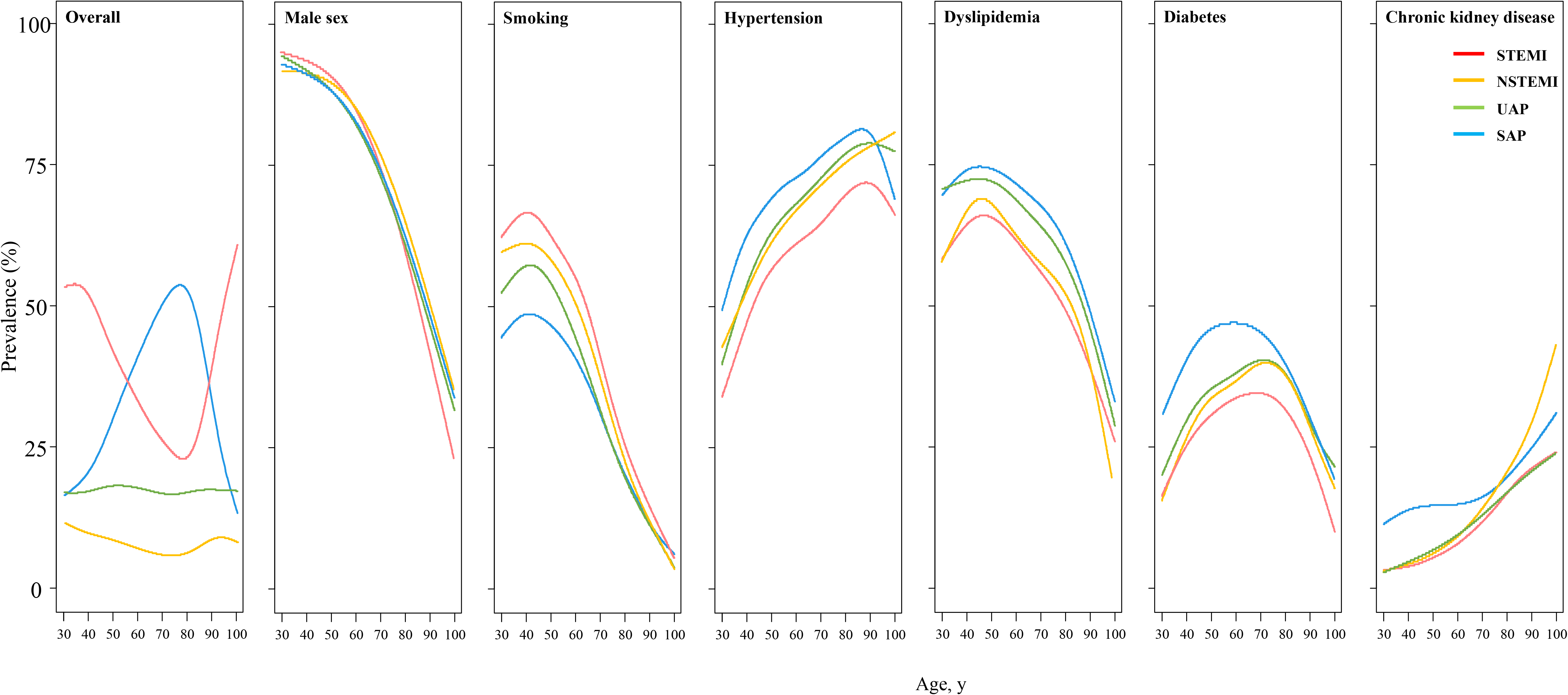
Age-specific prevalence of traditional atherosclerotic risk factors stratified by coronary artery disease type with a cubic spline.

Comparison of clinical profiles in the presence and absence of each traditional atherosclerotic risk factor is shown in **Table 2**. Compared with female patients, male patients were significantly younger (68.1 ± 11.6 versus 75.1 ± 10.2, P < 0.001) and more often smokers (40.4% versus 10.9%, P < 0.001). The smokers were also significantly younger (65.2 ± 11.6 versus 72.2 ± 11.0, P < 0.001) and less likely to have other risk factors. On the other hand, patients with CKD were significantly older (73.2 ± 10.8 versus 69.4 ± 11.7, P < 0.001), and more likely to have other comorbidities and critical conditions (cardiopulmonary arrest: 3.1% versus 2.2%, P < 0.001; acute heart failure: 9.5% versus 4.8%, P < 0.001; cardiogenic shock: 7.1% versus 3.9%, P < 0.001). Patients with dyslipidemia were less likely to present with critical conditions (cardiopulmonary arrest: 1.7% vs. 3.1%, P < 0.001; acute heart failure: 4.7% vs. 6.6%, P < 0.001; cardiogenic shock: 3.4% vs. 5.7%, P < 0.001). Patients with diabetes were more likely to have multivessel disease (56.4% vs. 66.0%, P < 0.001).

**Table 2.** Comparison of clinical profiles in the presence and absence of each traditional atherosclerotic risk factor.

|  | Overall<br>(N=559452) | Sex |  | Hypertension |  | Dyslipidemia |  | Diabetes |  | Smoking |  | Chronic kidney disease |  |
| --- | --- | --- | --- | --- | --- | --- | --- | --- | --- | --- | --- | --- | --- |
|  |  | Male<br>(N=410077) | Female<br>(N=149375) | Presence<br>(N=396021) | Absence<br>(N=163431) | Presence<br>(N=325887) | Absence<br>(N=233565) | Presence<br>(N=396021) | Absence<br>(N=347639) | Presence<br>(N=396021) | Absence<br>(N=377519) | Presence<br>(N=396021) | Absence<br>(N=478759) |
| Age, year | 70.0 ± 11.6 | 68.1 ± 11.6 | 75.1 ± 10.2 | 70.9 ± 11.2 | 67.8 ± 12.3 | 68.8 ±11.6 | 71.6 ± 11.6 | 69.6 ±11.0 | 70.1 ±12.0 | 65.2 ± 11.6 | 72.2 ± 11.0 | 73.2 ± 10.8 | 69.4 ± 11.7 |
| Male sex | 410077<br>(73.3%) | 410077<br>(100%) | 0<br>(0%) | 285192<br>(72.0%) | 124885<br>(76.4%) | 236719<br>(72.6%) | 173358<br>(74.2%) | 157300<br>(74.3%) | 252777<br>(72.7%) | 165597<br>(91.0%) | 244480<br>(64.8%) | 60024<br>(74.4%) | 350053<br>(73.1%) |
| Hypertension | 396021<br>(70.8%) | 285192<br>(69.6%) | 110829<br>(74.2%) | 396021<br>(100%) | 163431<br>(0%) | 247579<br>(76.0%) | 148442<br>(63.6%) | 161028<br>(76.0%) | 234993<br>(67.6%) | 124162<br>(68.3%) | 271859<br>(72.0%) | 64158<br>(79.5%) | 331863<br>(69.3%) |
| Dyslipidemia | 325887<br>(58.3%) | 236719<br>(57.7%) | 89168<br>(59.7%) | 247579<br>(62.5%) | 78308<br>(47.9%) | 325887<br>(100%) | 233565<br>(0%) | 133985<br>(63.3%) | 191902<br>(55.2%) | 112088<br>(61.6%) | 213799<br>(56.6%) | 44493<br>(55.1%) | 281394<br>(58.8%) |
| Diabetes | 211813<br>(37.9%) | 157300<br>(38.4%) | 54513<br>(36.5%) | 161028<br>(40.6%) | 50785<br>(31.7%) | 133985<br>(41.1%) | 77828<br>(33.3%) | 211813<br>(100%) | 347639<br>(0%) | 69688<br>(38.3%) | 142125<br>(37.7%) | 40904<br>(50.7%) | 170909<br>(35.7%) |
| Smoking | 181933<br>(32.5%) | 165597<br>(40.4%) | 16336<br>(10.9%) | 124162<br>(31.4%) | 57771<br>(35.4%) | 112088<br>(34.4%) | 69845<br>(29.9%) | 69688<br>(32.9%) | 112245<br>(32.3%) | 181933<br>(100%) | 377519<br>(0%) | 22660<br>(28.1%) | 159273<br>(33.3%) |
| Chronic kidney disease | 80693<br>(14.4%) | 60024<br>(14.6%) | 20669<br>(13.8%) | 64158<br>(16.2%) | 16535<br>(10.1%) | 44493<br>(13.7%) | 36200<br>(15.5%) | 40904<br>(19.3%) | 39789<br>(11.5%) | 22660<br>(12.5%) | 58033<br>(15.4%) | 80693<br>(100%) | 478759<br>(0%) |
| Prior heart failure | 44555<br>(8.0%) | 29354<br>(7.2%) | 15201<br>(10.2%) | 34822<br>(8.8%) | 9733<br>(6.0%) | 23417<br>(7.2%) | 21138<br>(9.1%) | 21699<br>(10.2%) | 22856<br>(6.6%) | 11747<br>(6.5%) | 32808<br>(8.7%) | 15146<br>(18.8%) | 29409<br>(6.1%) |
| COPD | 11827<br>(2.1%) | 10396<br>(2.5%) | 1431<br>(1.0%) | 8192<br>(2.1%) | 3635<br>(2.2%) | 6127<br>(1.9%) | 5700<br>(2.4%) | 4010<br>(1.9%) | 7817<br>(2.3%) | 5674<br>(3.1%) | 6153<br>(1.6%) | 2584<br>(3.2%) | 9243<br>(1.9%) |
| Peripheral artery disease | 32037<br>(5.7%) | 24097<br>(5.9%) | 7940<br>(5.3 %) | 25668<br>(6.5%) | 6369<br>(3.9%) | 18985<br>(5.8%) | 13052<br>(5.6%) | 16200<br>(7.7%) | 15837<br>(4.6%) | 10712<br>(5.9%) | 21325<br>(5.7%) | 10588<br>(13.1%) | 21449<br>(4.5%) |
| Myocardial infarction | 205449<br>(36.7%) | 154387<br>(37.7%) | 51062<br>(34.2%) | 134887<br>(34.1%) | 70562<br>(43.2%) | 112476<br>(34.5%) | 92973<br>(39.8%) | 68370<br>(32.3%) | 137079<br>(39.4%) | 80698<br>(44.4%) | 124751<br>(33.0%) | 25761<br>(31.9%) | 179688<br>(37.5%) |
| Number of vessels |  |  |  |  |  |  |  |  |  |  |  |  |  |
| 1 vessel | 348760<br>(62.3%) | 254144<br>(62.0%) | 94616<br>(63.3%) | 240748<br>(60.8%) | 108012<br>(66.1%) | 199328<br>(61.2%) | 149432<br>(64.0%) | 119476<br>(56.4%) | 229284<br>(66.0%) | 112576<br>(61.9%) | 236184<br>(62.6%) | 43362<br>(53.7%) | 305398<br>(63.8%) |
| 2 vessels | 140236<br>(25.1%) | 104168<br>(25.4%) | 36068<br>(24.2%) | 102602<br>(25.9%) | 37634<br>(23.0%) | 83891<br>(25.7%) | 56345<br>(24.1%) | 58103<br>(27.4%) | 82133<br>(23.6%) | 46354<br>(25.5%) | 93882<br>(24.9%) | 23172<br>(28.7%) | 117064<br>(24.5%) |
| 3 vessels | 70456<br>(12.6%) | 51765<br>(12.6%) | 18691<br>(12.5%) | 52671<br>(13.3%) | 17785<br>(10.9%) | 42668<br>(13.1%) | 27788<br>(11.9%) | 34234<br>(16.2%) | 36222<br>(10.4%) | 23003<br>(12.6%) | 47453<br>(12.6%) | 14159<br>(17.6%) | 56297<br>(11.8%) |
| Cardiopulmonary arrest | 12775<br>(2.3%) | 10161<br>(2.5%) | 2614<br>(1.8 %) | 7478<br>(1.9%) | 5297<br>(3.2%) | 5513<br>(1.7%) | 7262<br>(3.1%) | 4221<br>(2.0%) | 8554<br>(2.5%) | 4602<br>(2.5%) | 8173<br>(2.2%) | 2504<br>(3.1%) | 10271<br>(2.2%) |
| Acute heart failure | 30603<br>(5.5%) | 21211<br>(5.2%) | 9392<br>(6.3%) | 20991<br>(5.3%) | 9612<br>(5.9%) | 15219<br>(4.7%) | 15384<br>(6.6%) | 17825<br>(5.1%) | 12778<br>(6.0%) | 9795 ‡<br>(5.4%) | 20808<br>(5.5%) | 7650<br>(9.5%) | 22953<br>(4.8%) |
| Cardiogenic shock | 24437<br>(4.4%) | 17787<br>(4.3%) | 6650*<br>(4.5%) | 15159<br>(3.8%) | 9278<br>(5.7%) | 11149<br>(3.4%) | 13288<br>(5.7%) | 9051 †<br>(4.3%) | 15386<br>(4.4%) | 8292<br>(4.6%) | 16145<br>(4.3%) | 5712<br>(7.1%) | 18725<br>(3.9%) |

1 Continuous variables are given as mean ± standard deviation. Categorical variables are given as number (%). The comparison between presence and absent of each atherosclerotic risk factor are significant at P <0.001 unless
2 otherwise noted.
\*The comparison between presence and absent of each atherosclerotic risk factor are nonsignificant
† The comparison between presence and absent of each atherosclerotic risk factor are statistically significant at P <0.01
‡ The comparison between presence and absent of each atherosclerotic risk factor are statistically significant at P <0.05

### The strength of the association of TARFs with clinical profiles

As for the age of CAD onset requiring PCI, we evaluate the strength of association among TARFs by multiple regression model. The TARFs which associated with earlier age of CAD onset were smoking (unstandardized beta coefficient [UC], 2.68, 95% confidence interval [CI], 2.65-2.71, P<0.001), and male sex (UC, 2.67, 95% CI, 2.64-2.70, P<0.001). According to estimated explained relative risk model (R^2^ and RC), TARF most strongly associated with age at CAD onset was smoking (R^2^: 0.059/0.15, RC: 42.2%), followed by male sex (R^2^: 0.055/0.15, RC: 29.7%) (**Figure 4A <u>and</u> Figure 5**).

As for the presentation of multivessel disease, The TARFs which associated with multivessel disease were diabetes (UC, 0.068, 95% CI, 0.066-0.070, P<0.001), followed by CKD (UC: 0.066, 95% CI, 0.064-0.070, P<0.001) (**Figure 3B**). According to the relative risk model, TARFs most strongly associated with multivessel disease were diabetes (R^2^:0.10/0.17, RC: 59.4%) and CKD (R^2^:0.05/0.17, RC: 28.6%).(**Figure 4B <u>and</u> Figure 5**).

**Figure 3.**
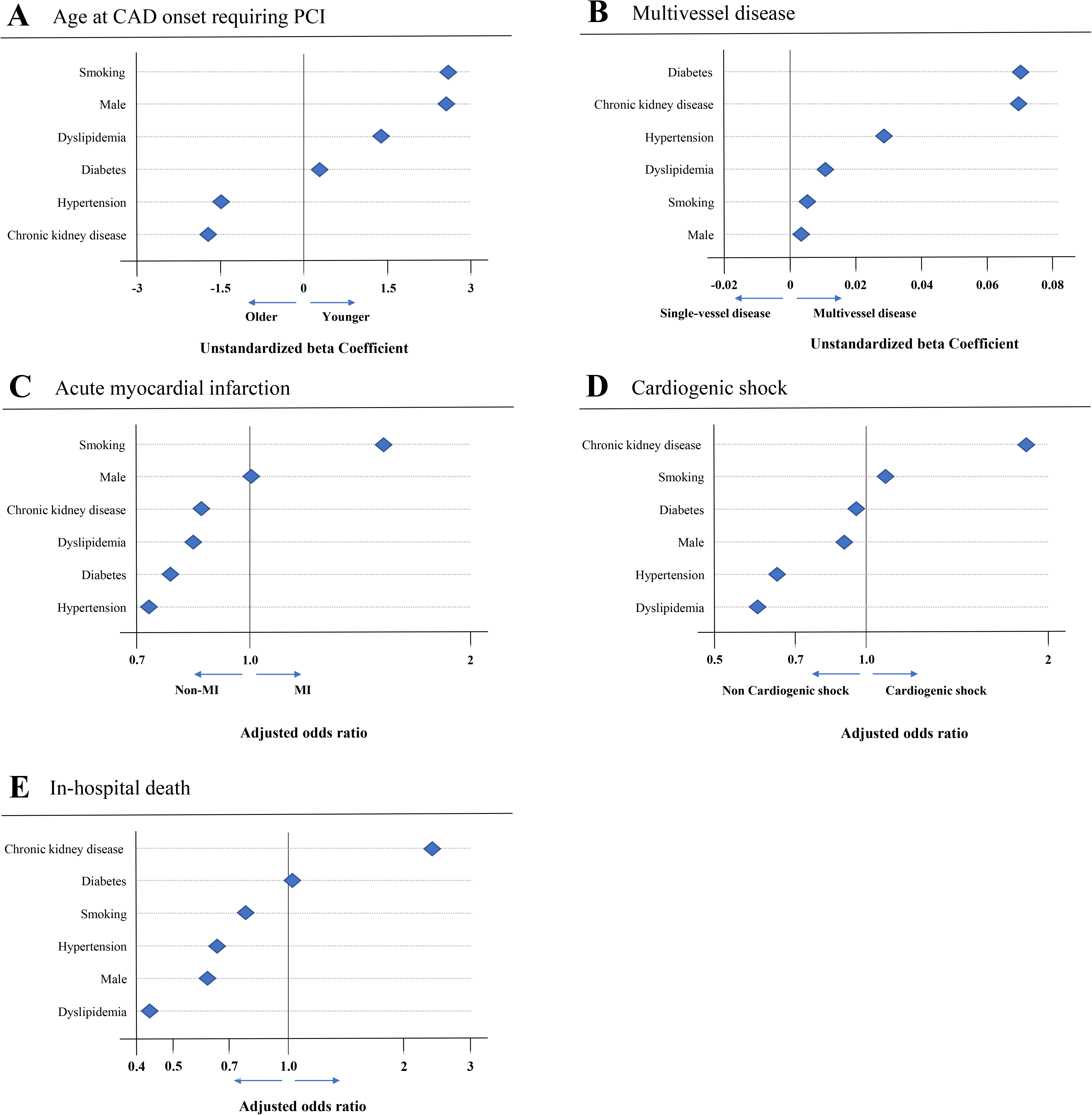
The unstandardized beta coefficient in multivariate multiple analysis or odds ratio in multivariate logistic analysis of traditional atherosclerotic risk factors for clinical profiles and mortality. (A) The unstandardized beta coefficient for the age of onset of coronary artery disease in multivariate multiple analysis. (B) The unstandardized beta coefficient for the number of vessel disease in multivariate multiple analysis. (C) The odds ratio for the presentation of myocardial infarction in multivariate logistic analysis. (D) The odds ratio for the presentation of cardiogenic shock in multivariate logistic analysis. € The odds ratio for in-hospital mortality in multivariate logistic analysis. Adjusted variables were as follows: hypertension, dyslipidemia, diabetes, smoking, and chronic kidney disease. CAD=coronary artery disease, PCI=percutaneous coronary intervention

As for the presentation of AMI, smokers (adjusted OR, 1.62, 95% CI, 1.60-1.64; P < 0.001) was more likely to develop AMI, whereas patients with diabetes (adjusted OR, 0.77, 95% CI, 0.76-0.78, P < 0.001) and those with hypertension (adjusted OR, 0.73, 95% CI, 0.72-0.74, P < 0.001) were less likely to develop AMI (**Figure 3C**). According to the relative risk model, the TARF most strongly associated with the presentation of AMI was smoking (R^2^:0.016/0.033, RC: 47.1%), while TARF most strongly associated with the presentation of non-AMI was diabetes (R^2^:0.007/0.033, RC: 21.9%) (**Figure 4C <u>and</u> Figure 5**).

**Figure 4.**
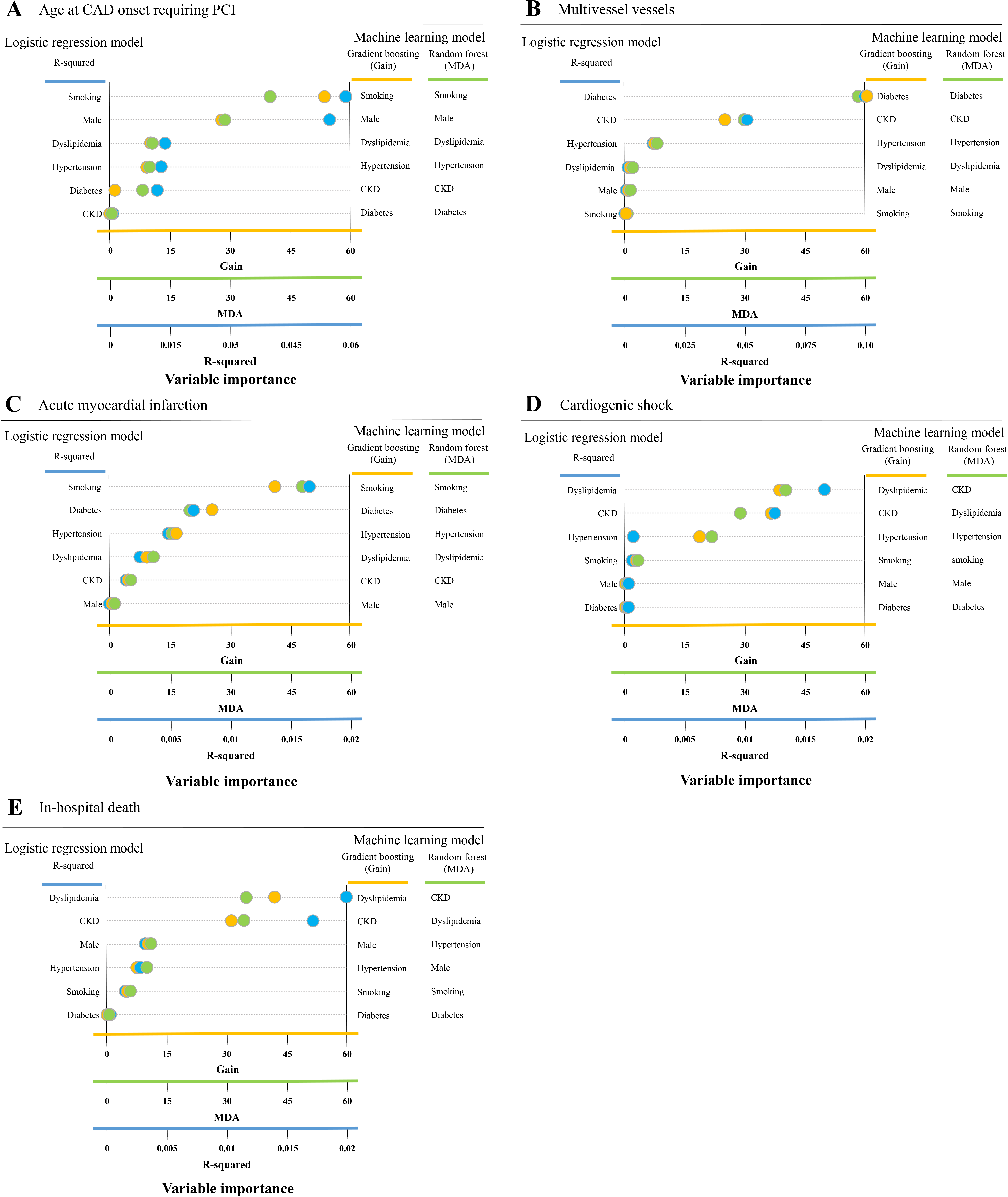
The relative importance of traditional atherosclerotic risk factors for clinical profiles and mortality of traditional atherosclerotic risk factors. The relative importance according to the R² value calculated by multiple regression analysis or logistics regression analysis, which is presented by the blue circle. The RI according to the gain calculated by gradient boosting is marked with the yellow circle. The RI according to the mean decrease accuracy calculated by random forest is presented by the green circle. (A) age at coronary artery disease onset, (B) multivessel disease, (C) presentation of acute myocardial infarction, (D) presentation of cardiogenic shock, and (E) in-hospital mortality. CAD=coronary artery disease, PCI=percutaneous coronary intervention

**Figure 5.**
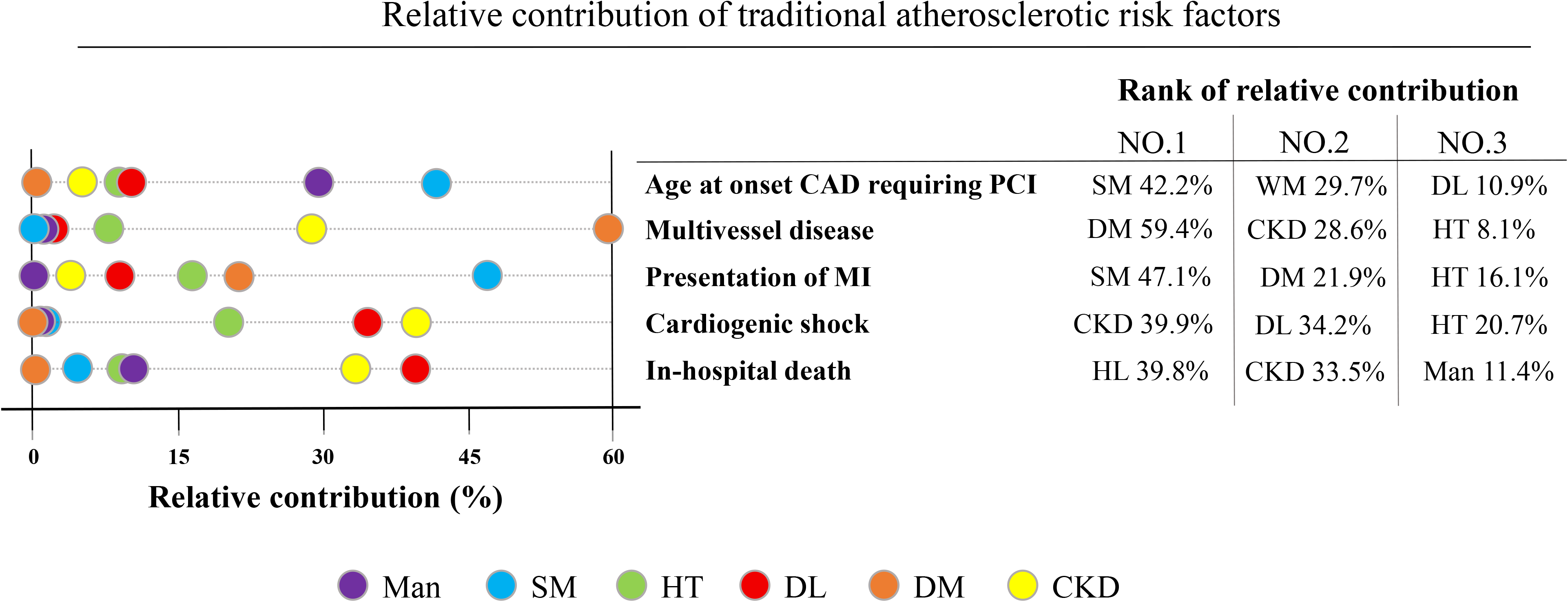
The relative contribution of each traditional atherosclerotic risk factor derived from all relative risk models for age at onset of coronary artery disease, multivessel disease, myocardial infarction, cardiogenic shock, and in-hospital mortality. The relative contribution was calculated by averaging the percentage of relative importance derived from R-squared, gradient boosting, and random forest. CAD=coronary artery disease, PCI=percutaneous coronary intervention, MS=male sex, SM=smoking, HT=hypertension, DL=dyslipidemia, DM=diabetes, CKD=chronic kidney disease

As for the presentation of cardiogenic shock, patients with CKD (adjusted OR, 1.95; 95% CI, 1.89-2.01, P < 0.001) had the highest risk, whereas the patients with dyslipidemia (adjusted OR, 0.62; 95% CI, 0.61-0.64, P < 0.001) and hypertension (adjusted OR, 0.68; 95% CI, 0.65-0.69, P < 0.001) had the lowest risk (**Figure 3D**). According to the relative risk model, CKD was most associated with cardiogenic shock status (R^2^:0.017/0.04, RC: 39.9%), followed by dyslipidemia (R^2^:0.013/0.04, RC: 34.2%) (**Figure 4D <u>and</u> Figure 5**).

### The strength of the association of TARFs with in-hospital mortality

CKD significantly increased the risk of in-hospital mortality (adjusted OR, 2.43; 95% CI, 2.29– 2.58; P < 0.001), whereas dyslipidemia significantly decreased the risk of in-hospital mortality (adjusted OR, 0.44, 95% CI, 0.41-0.46, P < 0.001) (**Figure 3E**). According to the relative risk model, the TARF with the strongest impact on in-hospital mortality was dyslipidemia. (R^2^: 0.021/0.049, RC: 39.8%), followed by CKD (R^2^:0.017/0.049, RC: 33.5%) (**Figure 4E <u>and</u> Figure 5**). Similarly, the protective effect of dyslipidemia on in-hospital mortality was observed in models that incorporated all risk factors (adjusted OR, 0.60; 95% CI, 0.57-0.64, P < 0.001, R^2^: 0.01/0.29, RC: 6.8%) (**<u>Figure S1</u>**).

### Age-adjusted and age-stratified strength of association of TARFs with clinical profile and in-hospital death

A comparison of clinical profiles between the presence and absence of each TARF by age stratification is shown in **<u>Table S1</u>**. The age-stratified impact of each TARF on multivessel disease, myocardial infarction, cardiogenic shock, and in-hospital death, and their age-specific impact were assessed by unstandardized coefficient, OR, and relative contribution (**<u>Table S2)</u>**. Although the distinctive effects of each TARF were observed in every age group, the degree of impact varied by age group. The associations between smoking and the presentation of AMI, diabetes and multivessel disease, dyslipidemia and cardiogenic shock and in-hospital death appeared to be more profound in the younger group. In addition, the same multivariate analysis and relative risk analysis was performed by adding age as a variable (**<u>Figure S2 and S3</u>**). Although age had the highest RI for in-hospital death, main result described above in the clinical profile was consistently observed.

## Discussion

In this report of 559,452 patients who underwent their initial PCI, we shed light on the differences in the prevalence and impact of each TARF on clinical profiles. In clinical practice, TARFs may be treated in parallel as a cause of atherosclerosis because they do not directly strongly affect mortality. To the best of our knowledge, this is the first study to examine the hypothesis with RI that patient conditions and clinical outcomes vary by TARFs. We also constructed the machine learning model (random forest), which takes into account complex higher-order interactions and provides robustness, bias reduction, and higher prediction accuracy ^16–19^.

This study showed that prevalence of TARF differed by CAD type and by age. While the prevalence of other TARFs remained low in patients with STEMI, this finding may be attributed to masking effects of STEMI during the acute phase, such as hypotension, lower LDL levels^20^, or stabilization of plaques by specific medications^21^. Conversely, the proportion of smokers in the patients with STEMI was significantly higher, and smoking was found to be one of the strongest predictors of AMI according to logistics analysis and relative risk model. Additionally, we observed that among premature CAD patients aged 30 to 40 years, approximately 60% had a history of smoking and dyslipidemia, and the proportion of males was significantly higher at 91%. smoking, male sex, and dyslipidemia accelerated the onset of CAD requiring PCI by approximately 7 years, 7 years, and 3 years, respectively. Dyslipidemia, which may onset during childhood due to genetic factors, may also contribute to the development of premature CAD ^22^. Since the prevalence of TARF varies with each age group, the TARF that should be actively intervened and educated for both primary and secondary prevention varies with each age group. Effective interventions to prevent premature CAD, which is increasing in incidence ^23^, should focus on smoking and dyslipidemia. Furthermore, our findings suggest that the plaque developed due to these risk factors is more likely to build up prematurely and may become prone to rupture compared to those associated with other TARFs. This difference could be explained due to the unique properties of atherosclerosis produced by each TARF and the host susceptibility or resilience to TARFs. Previous studies, which were small number, has shown that the properties of atherosclerosis varied by TARFs according to coronary computed tomography angiography and intravascular imaging device. Diabetes drives the development of multivessel disease and diffuse atherosclerotic plaque development more strongly than other TARFs^24^. Furthermore, in a separate study, plaques in smokers were found to be more strongly associated with high-risk plaque, such as low-attenuation plaques and Napkin-Ring-Sign, in coronary computed tomography angiography (CCTA) compared to other TARFs^25^. Similarly, gender difference in the atherosclerosis profile was identified in CCTA, which showed that the plaque in the male sex tends to be high-risk plaque^26^. Although TARFs are commonly associated with the development of atherosclerosis and the main biochemical mechanisms related to the development of atherosclerosis by each TARF have been outlined^14, 26–28^, the mechanisms underlying the differences in atherosclerosis profile and host susceptibility among TARFs remain to be elucidated. A better understanding of TARFs, which account for a large portion of atherosclerosis development, will further facilitate understanding and intervention in CAD. For example, to take more efficient preventive measures against smoking, which is unquestionably the largest contributor to premature CAD, if a susceptible population of young smokers could be identified, we can more rationally explain the importance of smoking cessation.

In this study, the strongest predictors of multivessel disease were diabetes, followed by and CKD. The tendency toward multivessel disease means that these risk factors are more likely to produce relatively stable plaques and diffuse atherosclerosis over time. In addition, the long-term lack of symptoms, which is often observed in patients with diabetes^29^, may have allowed the development of multivessel disease. Diabetes and age were the strongest predictors of SA in this study. The lack of difference in survival rates between older and newly diabetic patients and healthy controls in previous studies supports the idea that diabetes is unlikely to cause rapid progression of atherosclerosis or AMI^30^. The high RI of age for multivessel disease suggests that the duration of TARF may also be an important consideration.

Patients with dyslipidemia were less likely to have other comorbidities or critical conditions. Dyslipidemia was also strongly associated with cardiogenic shock and in-hospital mortality regardless of full model. In previous studies, the seemingly protective effect of dyslipidemia on mortality was consistent even after adjusting for various factors, including body mass index (BMI) as a correlation of dyslipidemia or medication for hyperlipidemia^31, 32^. However, since the analysis of association between TARFs and clinical profiles was cross-sectional analysis, it was not possible to differentiating cause and effect from simple association. ^33, 34^ In other words, it is impossible to discern from this study whether dyslipidemia is a cause or a consequence of shock. The benefits of statins may outweigh the risks of having a history of dyslipidemia, and the absence of dyslipidemia may reflect poor nutritional status or serious conditions, such as cancer^35^ and frailty^36^, that could not be adjusted for in our study. Patients with dyslipidemia often have hypertension, making it easier to introduce renin-angiotensin system inhibitors and other medications, which may have affected the mortality reduction. This finding is similar to the BMI paradox^37, 38^. Since this study included most patients who underwent PCI in Japan and was not a randomized controlled trial, which includes exclusion criteria and patients’ selection bias^39^, the interpretation of these results is only generalizable to patients who underwent PCI. Notably, these results do not apply to the general population. The correct interpretation of mortality related to dyslipidemia in this study is not that dyslipidemia reduces mortality, but that dyslipidemia is associated with a relatively lower risk of mortality than other TARFs in patients who underwent PCI. The mortality suppression effect of dyslipidemia was more significant in younger patients. The development of CAD at a younger age indicates a higher susceptibility of the host to the TARF. In addition, younger patients have fewer comorbidities, which purely reflect the influence of TARF, whereas older patients have numerous comorbidities, which creates complex interactions of various risk factors. Survival bias may also occur in the elderly.

## Limitations

The present study had several limitations. First, as mentioned above, the association between TARFs and clinical profile is observed in a cross-sectional study approach, so the causal relationship is uncertain. Although these association should be further evaluated in cohort studies, an enormous amount of effort and funding would be required considering the time to onset and incidence of premature CAD in the general population. Second, since most of the information in this study was not a continuous variable but a binary variable, the level-dependent impact of TARFs, such as blood pressure, serum creatinine level, serum lipids levels, level of hemoglobin A1c, and smoking amount per day, on CAD could not be estimated. Previous studies have shown that the severity of each TARF is important information regarding the outcomes. Furthermore, the impact of TARFs at levels that do not reach the diagnostic threshold has not been evaluated. Third, because the available variables were limited in this registry, unmeasured confounders, such as medications or detailed treatment, regarding mortality were present. Fourth, this result cannot be applied to CAD patients who did not undergo PCI. However, since most patients with MI (97.2%) underwent revascularization therapy in Japan^40^ and J-PCI registry includes approximately 85% to 90% of all PCI procedures in Japan, we presume that the generalizability of the present observations holds to some extent. Fifth, extremely old and young patients with PCI were not numerous, and this may have affected the results. Finally, the strength of the association between TARFs and long-term mortality remains unclear. As the course of atherosclerosis development depends on each TARF itself or individual susceptibility to TARFs, the importance of each risk factor for long-term mortality may vary in a time-dependent manner.

## Conclusion

The prevalence of each TARF varied with age and CAD type. Since the prevalence of TARF varies with each age group, the TARF that should be actively intervened and educated varies with each age group. Although TARFs have similar effects on the development of atherosclerosis, their impacts on the clinical profiles of CAD are not uniform, and their contribution to outcomes varies widely. These findings are expected to provide aid in the development of tailored interventions, and shared-decision making.

## Author Contributions

KK and KY designed the study. KY, SK, HI, KA, YI, and KK performed data analysis and drafted the manuscript. KK obtained the funding and supervised the study. All authors contributed to the interpretation of data and revision of the manuscript. KK is the guarantor.

## Data Availability

No additional data available.

## Acknowledgments

The authors would like to thank all the members of the CVIT and CVIT secretariat.

## Data availability statement

No additional data available.

## Ethical approval

As all data were anonymous and aggregated without any personal information, ethical approval was waived.

## Abbreviations and Acronyms

ACS: = Acute coronary syndrome
AMI: = Acute myocardial infarction
CAD: = Coronary artery disease
CCTA: = Coronary computed tomography angiography
CKD: = Chronic kidney disease
J-PCI: = The Japanese Percutaneous Coronary Intervention
OR: = Odds ratio
SA: = Stable angina
STEMI: = ST-elevated myocardial infarction
TARF: = Traditional atherosclerotic risk factor
UA: = Unstable angina
UC: = Unstandardized beta coefficient
PCI: = Percutaneous coronary intervention
RI: =Relative importance
RC: =Relative contribution

